# Plasma α-synuclein domain profiles across α-synucleinopathies

**DOI:** 10.1101/2023.07.17.23292753

**Authors:** Marie-Laure Pons, Pablo Mohaupt, Jérôme Vialaret, Etienne Mondesert, Margaux Vignon, Salomé Coppens, Moreau Stéphane, Sylvain Lehmann, Christophe Hirtz

## Abstract

**Background:** The differential diagnosis of α-synucleinopathies, including Parkinson’s disease (PD), dementia with Lewy bodies (DLB), and multiple system atrophy (MSA), is challenging due to overlapping clinical features and the current lack of reliable biomarkers. The primary diagnostic approach remains clinical, underscoring the need for objective biomarkers that can distinguish between these diseases. This study profiles α-synuclein peptides in plasma to explore potential disease-specific patterns.

**Methods:** We developed a targeted mass spectrometry-based assay to profile α-synuclein in plasma samples from PD (n=82), DLB (n=32), MSA (n=8), and controls (n=21). The assay quantifies non-modified peptides specifically derived from the N-terminus and NAC domain, regions implicated in aggregate formation, to assess potential disease-specific peptide patterns.

**Results:** No significant differences in peptide levels were observed between the disease groups, indicating consistent N-terminus and NAC domain profiles among α-synucleinopathies. However, a peptide within the NAC domain showed distinct patterns in MSA compared to other groups, which may reflect unique pathological processes.

**Conclusions:** This study provides the first blood-based assessment of α-synuclein peptide profiles, establishing a basis for future research into α-synucleinopathies. Refining the assay to include post-translationally modified peptides may enhance understanding of disease mechanisms and improve biomarker development.

## Background

Parkinson’s disease (PD) is a proteinopathy characterized by the misfolding and aggregation of α-synuclein into Lewy bodies and Lewy neurites in the brain parenchyma [1]. Similar pathological features are observed in dementia with Lewy bodies (DLB) and multiple system atrophy (MSA), with occasional co-occurrence in Alzheimer’s disease (AD) [2]. The primary diagnostic approach for these conditions remains clinical; DLB is diagnosed based on the McKeith criteria, which stipulate that cognitive dysfunction should precede or occur concurrently with motor dysfunction [3]. In contrast, PD typically manifests initially with motor symptoms and may later progress to cognitive deficits, termed PD with dementia (PDD) [4]. MSA is uniquely characterized by α-synuclein aggregates in oligodendroglial cells, and is diagnosed primarily through clinical symptoms and MRI findings that aid in differentiation from other α-synucleinopathies [5]. The differential diagnosis among these disorders is hampered by overlapping clinical symptoms, particularly in the early stages of PD and MSA. A key pathogenic mechanism in α-synucleinopathies, known as ‘seeding’, involves the recruitment of soluble α-synuclein monomers by misfolded α-synuclein fibrils, promoting their aggregation into neurotoxic amyloid structures [6]. Recent advancements in seed amplification assays have shown promise in distinguishing these disorders by detecting distinct patterns of α-synuclein aggregation in cerebrospinal fluid (CSF) [7–13]. Further research suggests that the diagnostic value of these assays can be enhanced by combining them with measurements of neurofilament light chain (NfL) [14,15]. These developments indicate a potential shift from a purely clinical to a more biologically oriented diagnostic approach [16].

With the advancement of disease-modifying therapies, accurate and efficient patient stratification is essential for targeted treatments. The close resemblance among α-synucleinopathies necessitates biomarkers capable of detecting subtle differences in disease mechanisms and progression, thereby enabling precise patient classification and monitoring throughout therapeutic interventions. Distinguishing between α-synucleinopathies is biologically challenging due to the similar Lewy body structures observed in PD and DLB [17]. We hypothesize that fluid biomarkers in α-synucleinopathies may result from distinct protease activities generating unique truncated α-synuclein forms or from disease-specific post-translational modifications. Truncation typically affect the C-terminus, with the N-terminal and the non-amyloid-β component (NAC) domains remaining intact within Lewy bodies. While the specific proteases involved are unknown, further studies are needed to determine if variations in these truncated forms are disease-specific, and how they are reflected in biofluids. In α-synucleinopathies, α-synuclein is consistently phosphorylated at the 129th amino acid residue, a modification detectable in skin biopsies with 93% accuracy for identifying these conditions [18]. This phosphorylation is present across all α-synucleinopathies, indicating it is not disease-specific. In contrast, phosphorylation at amino acid residue 64 is unique to PD and absent in MSA, suggesting it could differentiate these disorders [19]. We developed a mass spectrometry-based approach to analyze plasma α-synuclein levels and domain distributions, aiming to identify biomarker signatures that could improve diagnostic accuracy and patient stratification.

## Methods

### Study participants

Participants were recruited at the Montpellier Memory Resources Center (CMRR). The study cohort included 143 individuals with clinically diagnosed PD (n=82), DLB (n=32) and MSA (n=8) and 21 control subjects with non-neurodegenerative conditions. These conditions included neurologic affections such as neuropathic (23.8%), vascular (28.6%), immunologic (28.6%), and hydrocephalus (19.0%). Diagnosis of PD and MSA were established according to the International Parkinson and Movement Disorder Society (MDS) criteria, while DLB was diagnosed based on the McKeith criteria [3,20]. Plasma samples were collected and stored at the Montpellier Neurobank (CHU resource center BB-0033-00031). Ethical approval was obtained from the Montpellier University Hospital’s regional Ethics Committee, and written informed consent was obtained from all participants. The demographic and clinical characteristics of participants are summarized in Table 1.

**Table 1.** Demographic characteristics of the patient cohorts.

| Demographic<br>characteristic | PD | DLB | MSA | Controls |
| --- | --- | --- | --- | --- |
| N | 82 | 32 | 8 | 21 |
| Age at sampling (years) | 73.5<br>[ 67 – 77.75 ] | 70<br>[ 66.75 – 75.33 ] | 71.6<br>[ 66.65 – 76 ] | 68<br>[ 65 – 74.7 ] |
| Sex (male/female) | 50 / 32 | 27 / 5 | 3 / 5 | 16 / 5 |
| Disease duration<br>(years) | 7.5 [ 3 – 12 ]<br>n = 78 | 3 [ 1 – 4 ]<br>n = 31 | 4.2 [ 3 – 7.4 ] | 1.8 [ 0.5 – 3 ] |
| Cognitive score<br>(MMSE, MOCA or<br>MDRS) % |  |  |  |  |
| Positive | 30.5 | 50.0 | 9.4 | 9.5 |
| Negative | 17.1 | 37.5 | 87.5 | 52.4 |
| N/A | 52.4 | 12.5 | 3.1 | 38.1 |
| Hoehn and Yahr stage<br>(UPDRS 5) % | Median<br>Stage: 3<br>(32.9%), Main<br>Range: Stages<br>2-3 (78%) | N/A | N/A | N/A |

Note: Demographic characteristics are presented as medians and interquartile ranges. For categorical variables, differences across groups were evaluated using the chi-square goodness of fit test, and for continuous variables, the Kruskal-Wallis test was employed. The cognitive score is presented as positive or negative using cut-off values of 27 for MMSE, 24 for MOCA and 130 for MDRS

### Sample Preparation

EDTA plasma samples (95 µL) were thawed on ice for 1 hour and diluted with 855 µL of deionized water. Samples were supplemented with 5.7 µL of 10 ng/µL recombinant full-length α-synuclein uniformly labeled with 15N (LGC, Teddington, UK) in 50 mM ammonium bicarbonate. After vortexing, 142.5 µL of 70% perchloric acid was added to precipitate non-target proteins, followed by a 15-minute incubation on ice. The samples were centrifuged at 16,000 xg for 15 minutes at 4°C. Supernatants were transferred to LoBind tubes, and 95 µL of 1% trifluoroacetic acid was added. The samples were concentrated at room temperature using a vacuum concentrator (Speedvac, Labconco). Solid-phase extraction (SPE) was performed using RP-W tips on an AssayMap Bravo (Agilent Technologies). The tips were first equilibrated with water before loading the samples. After sample loading, the tips were washed with 10% acetonitrile containing 0.1% formic acid. The proteins of interest were then eluted with 45% acetonitrile containing 0.1% formic acid. The eluates were dried in a vacuum concentrator for 90 minutes at room temperature and reconstituted in 20 µL of 50 mM ammonium bicarbonate. Digestion was performed by adding 7 µL of 1 µg/µL Trypsin/LysC, followed by a 4-hour incubation at 37°C with gentle agitation (450 rpm). Digestion was stopped by adding 0.5 µL of formic acid.

### Liquid chromatography-mass spectrometry multiple reaction monitoring analysis

Sample were analyzed using a liquid chromatography system (Mikros, Shimadzu Corporation) coupled to an LCMS-8060 triple quadrupole mass spectrometer (Shimadzu Corporation) in positive ionization mode. Chromatographic separation was performed on a ZORBAX SB-Aq reversed-phase column (1 × 150 mm, particle size 3.5 µm, Agilent Technologies), at 35°C. The mobile phases were 0.1% formic acid in LCMS-grade water (phase A) and 0.1% formic acid in acetonitrile (phase B), with a 30-minute gradient increasing phase B from 0% to 30% at a flow rate of 50 µL/min. Multiple Reaction Monitoring (MRM) method was employed with scheduled retention time windows. The ion source settings were optimized, including a nebulizing gas flow rate of 3 L/min, heating gas flow rate of 10 L/min, interface temperature of 300°C, desolvation line temperature of 250°C, heating block temperature of 400°, and drying gas flow rate of 10 L/min. Collision energy and dwell time were individually optimized for each peptide. Details on peptide positions and transitions are provided in Table 2.

**Table 2.**
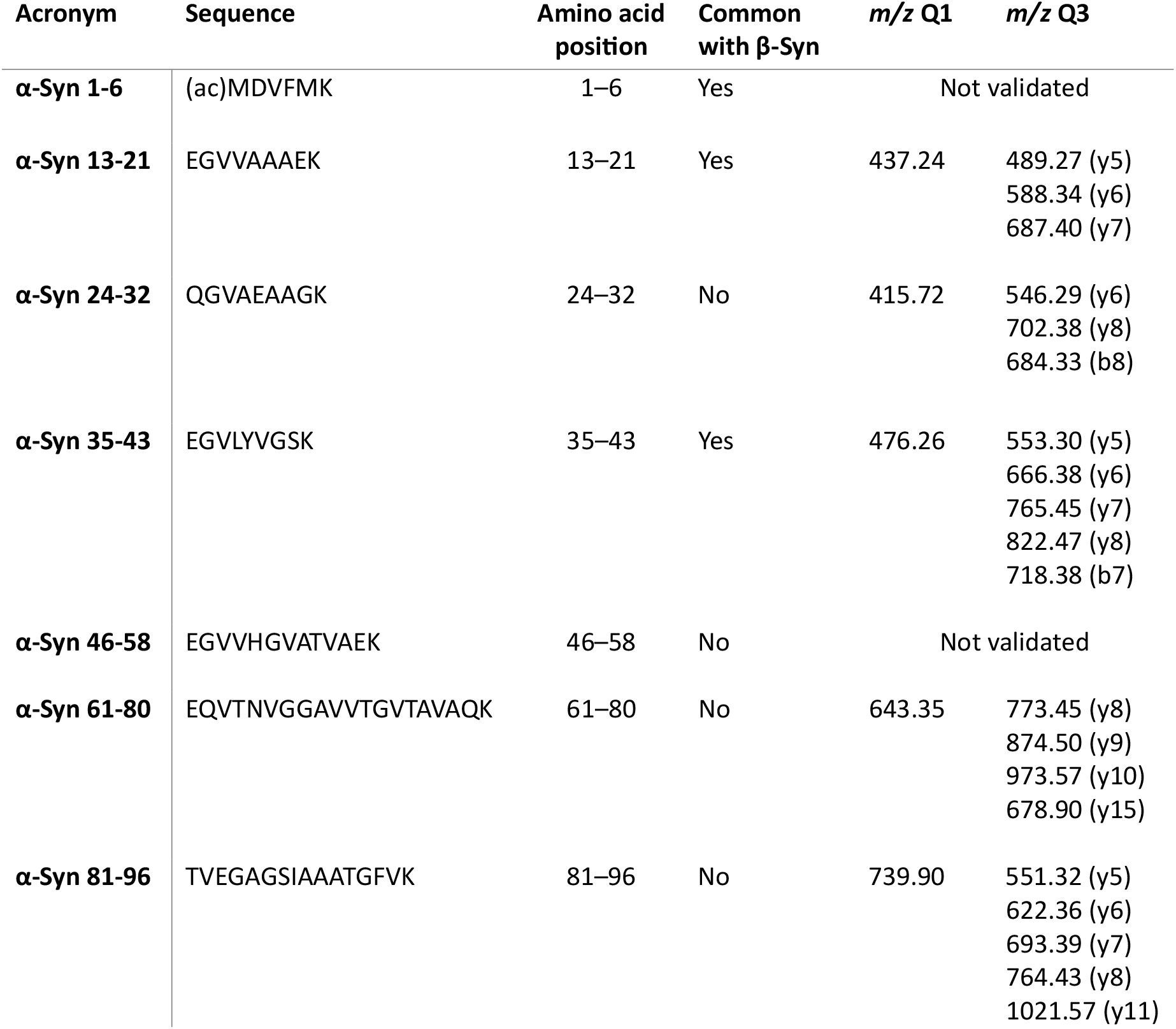
Selected α-synuclein peptides and their respective amino acid position, mass to charge ratio monitored in Q1, and fragment ions monitored in Q3.

Plasma QC samples with low (46.11 ng/mL), mid (74.70 ng/mL), and high (140.24 ng/mL) concentrations of α-Synuclein were used for method validation. The validation included assessment of intra-assay coefficients of variation (CV) at low (4.9-13%), mid (3.3-8%), and high (2.5-6%) concentrations, and inter-assay CVs at low (12-22%), mid (13-16%), and high (10-14%) concentrations.

Sample stability was evaluated on ice (6 hours, recovery 88 – 103%), at room temperature (4 hours, recovery 99 – 106%), and in the autosampler (48 hours, recovery 95 – 121%). Dilution linearity was confirmed up to a four-fold dilution with accuracy ranging from 87 – 117%, except for α-Syn 35-43 and α-Syn 81-96, which showed accurate results up to a two-fold dilution (87 – 94%). Parallelism was tested to a four-fold dilution with accuracy between 86 – 100%, except for α-Syn 61-80 and α-Syn 81-96, which were accurate to a two-fold dilution (106 – 119%). The intra-assay CV during patient sample measurement, as determined using plasma QC sample, was 20.3 – 29.8% for α-synuclein-specific peptides and 18.7% and 40.0% for peptides shared between α-synuclein and β-Synuclein, specifically α-Syn 13-21 and α-Syn 35-43, respectively.

### α-Synuclein quantification by commercially available Immunoassay

Plasma samples were analyzed using an immunochemiluminescence assay (MesoScale Discovery) to quantify total α-synuclein concentrations. Samples were diluted 200-fold prior to analysis, and all measurements were performed in accordance with the manufacturer’s instructions. The capture antibody was a rabbit monoclonal antibody targeting the C-terminal region of α-synuclein (amino acid residues 110-125), while the detection antibody was a mouse monoclonal antibody targeting amino acid residues 15-125.

### Data processing and statistical analysis

Data were processed using Skyline Software (version 20.1.0) and LabSolutions Insight Browser. Peaks were visually inspected and adjusted for accurate peak area calculations. Peptide abundances were quantified by comparing endogenous peptides with isotopically-labeled standards.

Statistical analyses were performed in R (version 4.3.0). Homogeneity of variances was assessed with Levene’s test, and normality was evaluated using Shapiro-Wilk test, indicating deviations from normality on both raw and log-transformed residuals. Group comparisons were made using rank-based ANCOVA, adjusting for age and sex. Peptide profiles were visualized using median abundance and ridgeline plots. Correlations between peptides were assessed with Spearman’s rank correlation coefficients, visualized with correlograms. To assess the differential regulation among groups, receiver operating characteristic (ROC) curves were generated from logistic regression models that evaluated the contributions of peptides without incorporating confounders.

## Results

### Group-wise comparison at the peptide-level for differential diagnosis in plasma

A group-wise comparison at the peptide level reveals similar profiles across conditions (Figure 1). Specifically, our data suggest that α-synuclein levels may be marginally elevated in DLB compared to controls and slightly higher than in PD. However, these differences were not statistically significant (p > 0.05). The modest elevation observed in MSA likely reflects the small sample size (n = 8) and also did not reach statistical significance (p > 0.05). The comparable levels of α-Syn 13-21 and α-Syn 35-43 with the other peptides suggest a limited impact of β-synuclein, given that α-Syn 13-21 and α-Syn 35-43 are also present in this protein. Plasma β-synuclein levels could not be included as a confounding factor due to the limited availability of blood-based assays for this protein [21]. ELISA assay results, which measure total α-synuclein, did not show significant differences among groups. Furthermore, the concentrations measured by the ELISA vary widely among patient samples. This may be attributed to the choice of capture antibody that targets the C-terminus, an area known to undergo truncation and post-translational modification. There was no correlation observed between measurements and UPDRS scores.

**Figure 1.**
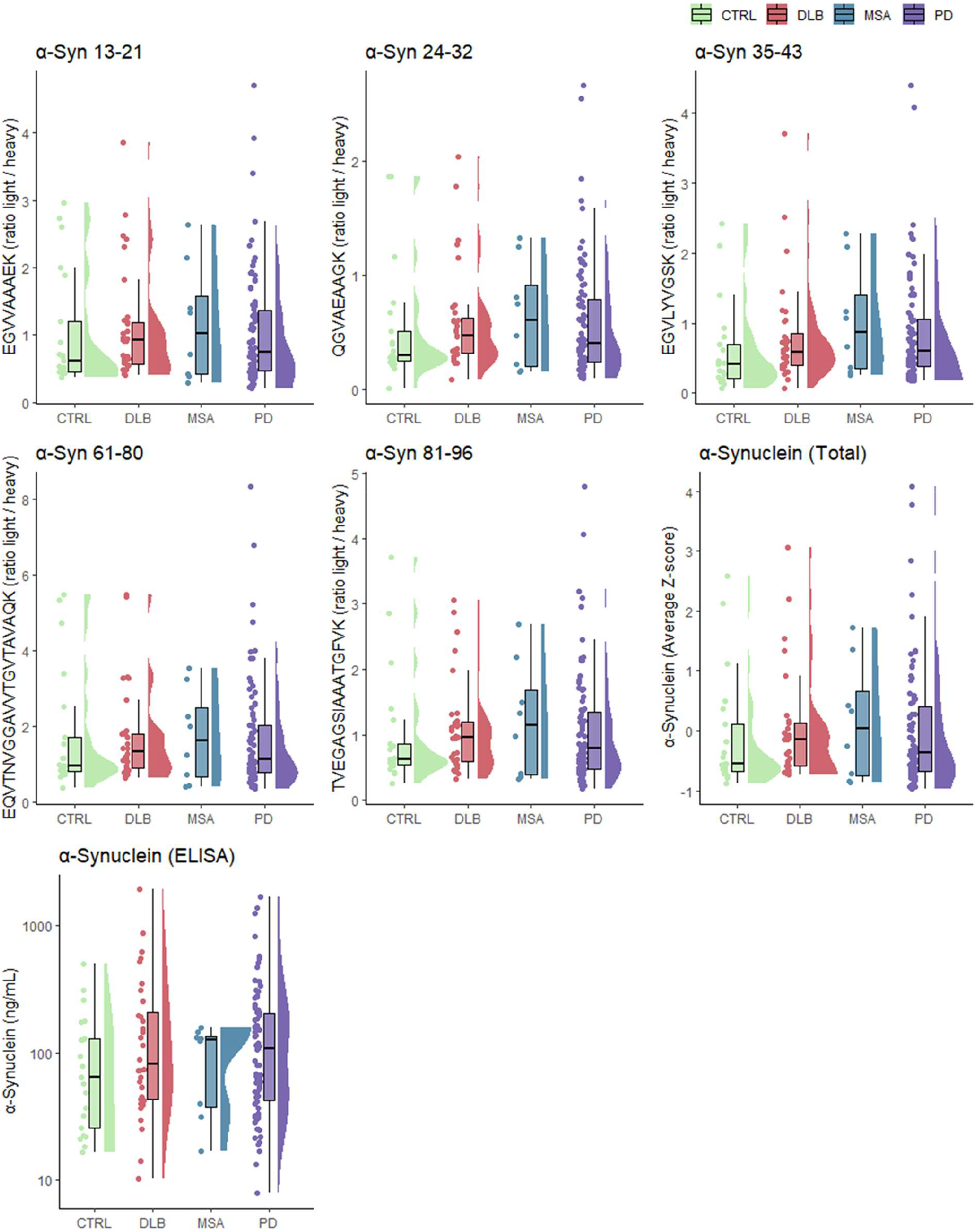
Group-wise comparison of α-Synuclein and corresponding peptide levels in clinical samples. Comparative analysis of α-Synuclein peptides originating from the N-terminus and the NAC-domain in plasma samples obtained from participants with dementia with Lewy bodies (DLB, n = 31), multiple system atrophy (MSA, n = 8), Parkinson’s disease (PD, n = 82), and controls (CTRL, n = 21). The analysis includes individual boxplots for each peptide, a combined boxplot with mean Z-scored peptides representing total α-Synuclein, and a boxplot of ELISA results for α-Synuclein. Statistical comparisons were performed using rank-based ANCOVA with age and sex as covariates, followed by Bonferroni’s post hoc test. The boxplots display the median, interquartile range, and 1.5 interquartile range as a horizontal line, box, and whiskers, respectively. The half violin plots illustrate data distribution. Each dot represents an individual sample measurement. Significance labels (*P ≤ 0.05, **P ≤ 0.01, ***P ≤ 0.001, ****P ≤ 0.0001) would be indicated if applicable.

### Profiling plasma α-synuclein peptides in clinical samples

We analyzed the peptide abundances across various disease groups to understand central tendencies and patterns (Figure 2A). Our findings again indicate a slight increase in α-synuclein levels in α-synucleinopathies compared to controls. This increase is consistent along the entire N-terminus and NAC domain. Additionally, the patterns of peptide abundance are remarkably similar among the different disease groups, suggesting a conserved trend among these peptides. To further explore the distribution of peptide abundances within each disease group, we generated ridgeline plots (Figure 2B). These plots provide a visual representation of the density distribution of peptide abundances, allowing for a detailed comparison of peptides within each disease group. While the ridgeline plots do not directly highlight increased α-synuclein levels, they reveal the overall distribution patterns and variability of peptide abundances across the disease groups, complementing the findings from the median abundance plot. The broader density distribution observed in MSA most likely represents the small group size.

**Figure 2.**
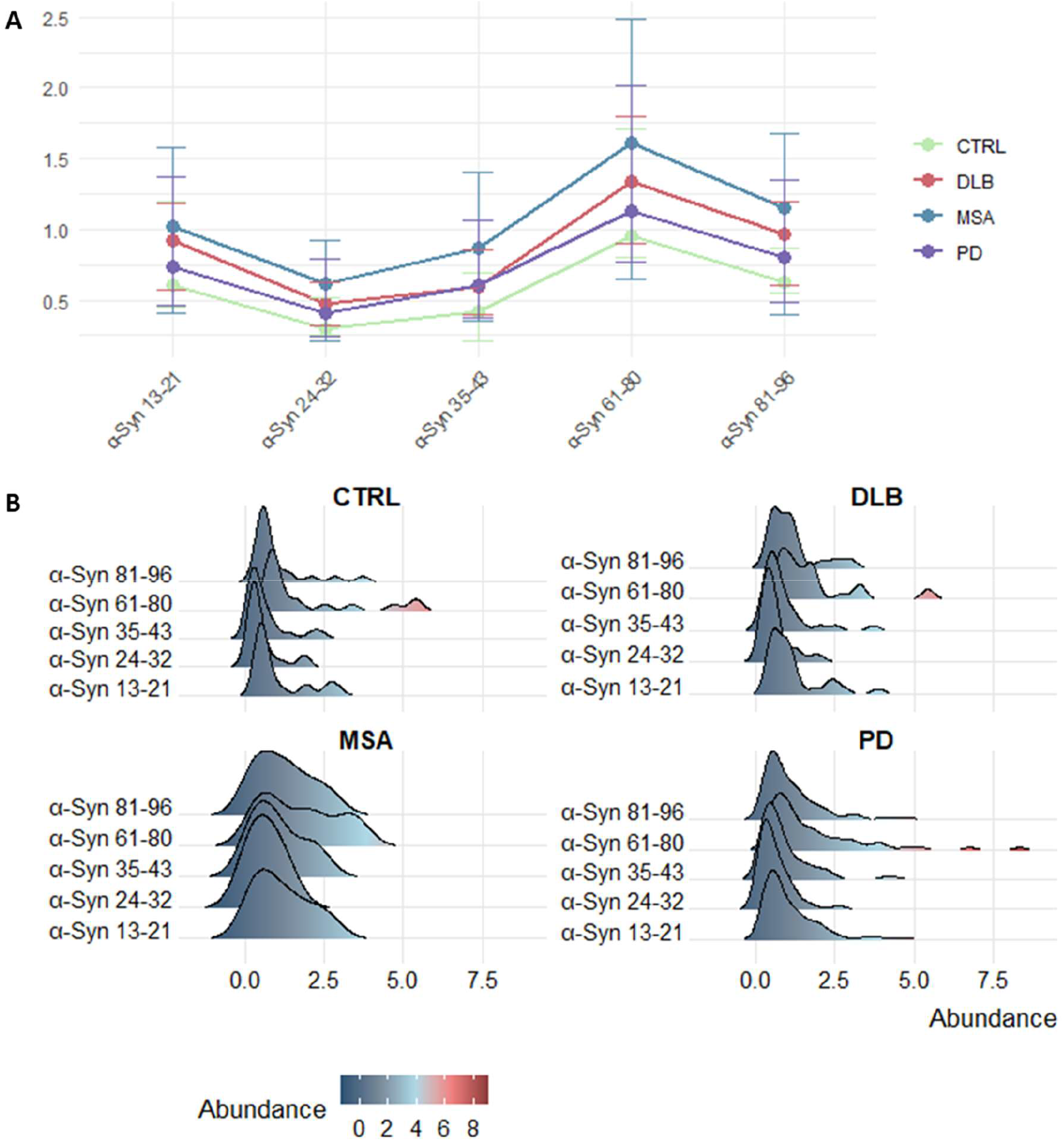
Profiling plasma α-synuclein peptides in clinical samples. (A) This figure displays the median abundance of α-synuclein peptides in plasma in different disease groups, including Parkinson’s Disease (PD), Control (CTRL), Dementia with Lewy Bodies (DLB), and Multiple System Atrophy (MSA). Each peptide is ordered along the x-axis by their amino acid position. The median abundances are depicted as colored points, with error bars representing the interquartile ranges (IQR). The disease groups are color-coded: CTRL (light green), DLB (red), MSA (blue), and PD (purple). Lines connect the median values for each disease group across the different peptides to facilitate comparison. (B) Presents the distribution of α-synuclein peptide abundances across the same disease groups. Each panel represents a different disease group, and within each panel, the abundance distributions of various peptides are shown using ridgeline plots. The peptides are ordered along the y-axis by their amino acid position. The ridgeline plots display the density of peptide abundances, with colors indicating the abundance levels. The color gradient ranges from dark blue for low abundances to dark red for high abundances. The x-axis represents the abundance levels, and the y-axis lists the peptides.

### Associations between plasma α-synuclein peptides

To assess whether peptide levels were interrelated within each diagnostic group, correlograms were utilized to visualize the associations (Figure 3). These correlograms revealed strong correlations between peptides across all diagnostic groups, with Spearman’s rho ranging between 0.71 and 1.00, and p-values < 0.001. Associations with the ELISA were generally weak, with significant correlations only observed in the PD group (p < 0.01 and p < 0.001), likely due to the larger sample size in this disease group. The positive correlations between peptides indicate a synchronous elevation across peptide levels. This finding, however, does not exclude the possibility of disproportionate regulation across different regions, as peptide levels can correlate while exhibiting disproportionate increases within a specific group.

**Figure 3.**
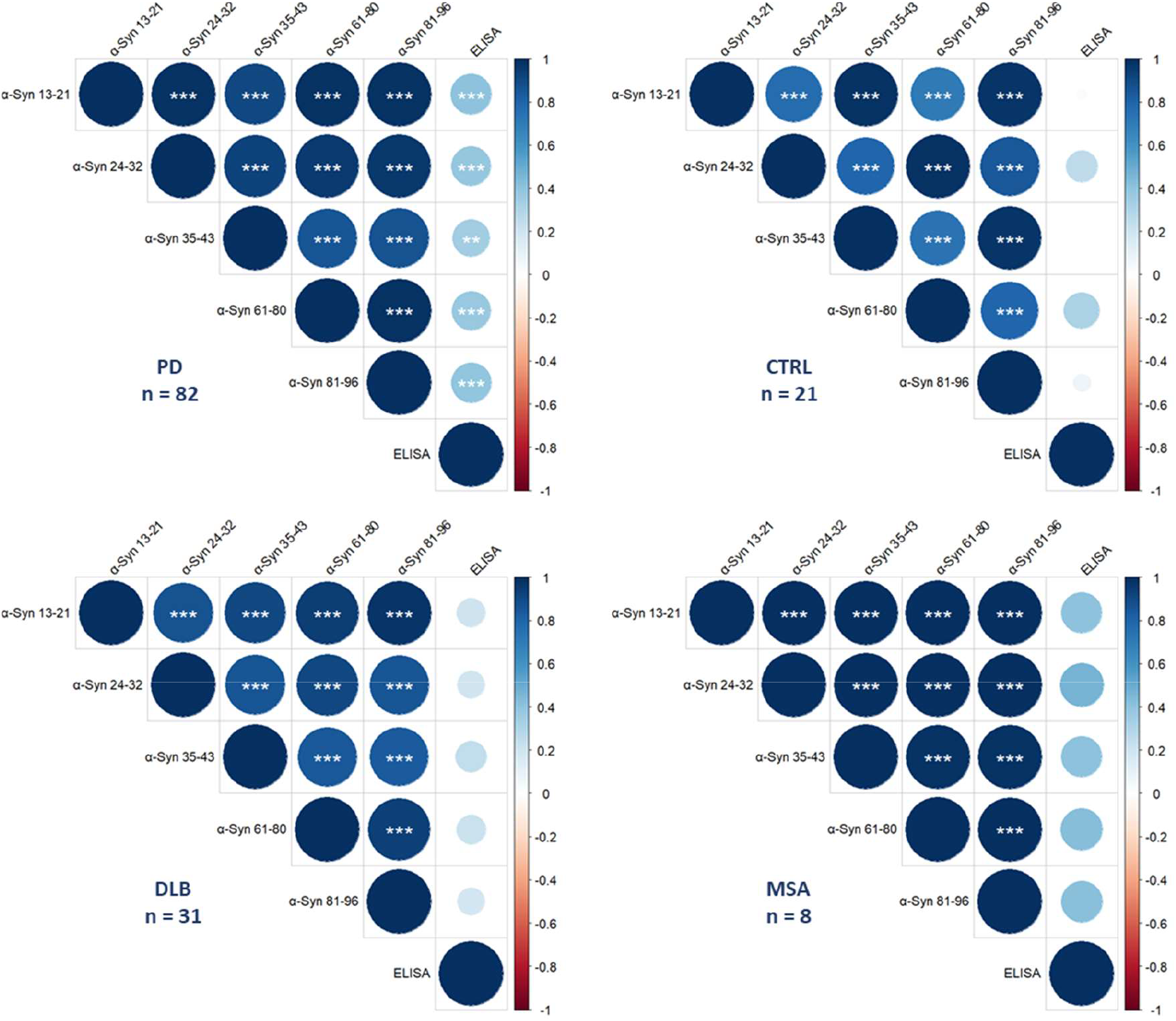
Correlogram depicting α-synuclein peptide correlations in plasma within each diagnostic groups. The correlation matrices, visualized as correlograms, are based on Spearman’s rank correlation coefficients. Circle size is proportional to the absolute value of the correlation coefficient, with larger circles indicating stronger correlations. Circle color indicates the direction of the correlation: positive correlations (blue) suggest that as one variable increases, so does the other, while negative correlations (red) indicate that as one variable increases, the other decreases. Darker shades represent stronger correlations. Significance levels are indicated as follows: *P ≤ 0.05, **P ≤ 0.01, ***P ≤ 0.001.

### Exploring differential regulation through classification based on plasma α-synuclein peptide combinations using logistic regression

To evaluate whether peptides were differentially regulated in specific groups, we conducted ROC analysis by creating peptide combinations using logistic regression models (Figure 4, Supplementary material 1). Notably, models including α-Syn 35-43 showed higher areas under the curve (AUCs). However, this is likely due to the high %CV during the cohort, but values are still provided for reference and interpretation. The resulting AUCs and Youden’s indices, as presented in Table 2, do not directly indicate aberrant regulation among peptides. Specifically, no differences were observed between PD and DLB, with AUCs not exceeding 0.60 and Youden’s indices < 0.25. Furthermore, PD and DLB did not show discernible aberrations compared to control samples, with AUCs not exceeding 0.68 and Youden indices < 0.40. The MSA group showed potential dysregulation in the NAC-domain, with combinations of α-Syn 61-80 and α-Syn 81-96 reaching AUCs of 0.71, 0.68, and 0.74 and Youden’s indices of 0.5, 0.44, and 0.54 against PD, DLB, and controls, respectively. Other combinations with α-Syn 61-80 also showed higher AUCs, potentially indicating some aberrant regulation of this peptide. However, it should be noted that this effect might be due to disproportionate group sizes.

**Figure 4.**
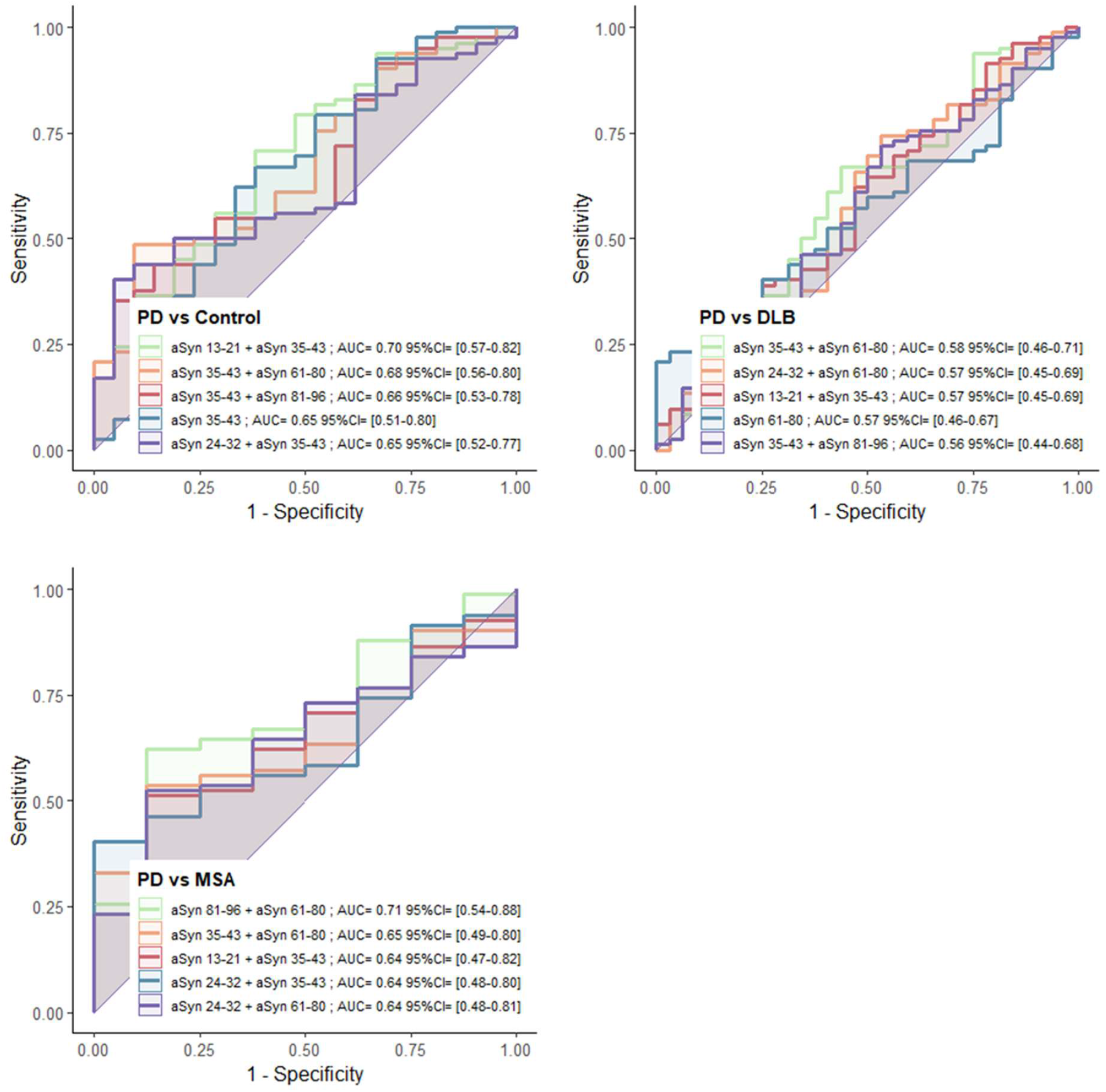
Receiver Operating Characteristic (ROC) curves for classification based on α-synuclein peptide combinations. ROC curves illustrate the diagnostic accuracy of individual peptides and peptide combinations derived from logistic regression models. The curves depict the ability to differentiate Parkinson’s disease (PD) from dementia with Lewy bodies (DLB), multiple system atrophy (MSA), and control groups. Highlighted are the five peptides or peptide combinations with the highest area under the curve (AUC) values, indicating the most significant diagnostic performance.

## Discussion

In this study, we developed a novel mass spectrometry-based assay to monitor peptides from the N-terminus and NAC domain of α-synuclein in plasma, marking the first assessment of peptide levels from distinct regions of α-synuclein in blood. We hypothesized that disease-specific truncations or post-translational modifications would alter levels of non-modified α-synuclein peptides across α-synucleinopathies. However, no significant differences were observed between disease groups, suggesting that N-terminus and NAC domain profiles remain consistent, with slight elevations in α-synucleinopathies compared to controls. This uniformity may reflect shared mechanisms or indicate that disease-specific differences are subtle and not detected by non-modified peptide measurements.

α-Synucleinopathies share pathogenic features with tauopathies, such as the aggregation of misfolded proteins into toxic fibrils, suggesting that advances in AD diagnostics may inform biomarker development for similar proteinopathies. In AD, biomarkers like phosphorylation of tau at the 217th amino acid (pTau-217) highlight the role of post-translational modifications in distinguishing AD from other tauopathies, while N-terminal tau, linked to truncation, illustrates how targeting specific protein regions reflects disease mechanisms [22–27]. The Aβ42/40 ratio further exemplifies this, capturing the differential cleavage of amyloid precursor protein to produce aggregation-prone Aβ42, whose reduction in biofluids serves as a key marker of amyloid pathology. These findings underscore the importance of monitoring specific protein regions and modifications to gain insights into disease mechanisms and guide the development of targeted biomarkers.

Our study explored whether similar principles observed in AD apply to α-synucleinopathies, focusing on potential disease-specific truncation or post-translational modifications of α-synuclein. Although we found no significant disproportional regulation at the peptide level, comparisons between PD and MSA involving α-synuclein peptide 61-80 suggested aberrant regulation. This effect was not linked to known phosphorylation at the 64th amino acid residue in PD, as it was not observed when comparing PD with controls [19]. The same pattern was observed in comparisons of MSA with DLB and controls. This peptide, part of the NAC region and identified as a key contributor to the formation of aggregates suggests alternative regulatory mechanisms in MSA that may reflect unique pathological features [28]. However, these findings require cautious interpretation due to small and disproportionate group sizes, which can affect the reliability and robustness of ROC analysis. Moreover, our focus on non-modified peptides does not exclude the possibility of differential regulation through disease-specific truncations or modifications occurring at levels that minimally affect non-modified peptide measurements.

This study contributes to the expanding research on α-synucleinopathies to improve diagnostic capabilities. There is an urgent clinical need for accessible biomarkers that can differentiate between PD and MSA during the early stages of these conditions. As disease-modifying therapies advance, there remains a critical need for reliable methods to monitor disease progression. This need may not be adequately met by current seed amplification assays, highlighting the importance of developing new diagnostic biomarkers and a better understanding of α-synucleinopathies.

### Limitations

Our study employed an unconventional approach by investigating the disproportionate representation of peptides in plasma rather than CSF, which is preferred for such studies as it more accurately reflects brain pathologies *in vivo*. Therefore, effects observable in CSF may be missed or appear diluted in our results due to the peripheral expression of α-synuclein. Additionally, the small sample size of the MSA group and the disproportional group sizes could skew the results, particularly in ROC analysis.

Consequently, any conclusions regarding MSA should be interpreted with caution and require validation in subsequent studies. Furthermore, these initial findings specifically monitor non-modified peptides, whereas a more comprehensive approach would also target their post-translationally modified forms. The increased %CVs during patient sample analysis also limit the accuracy of our outcomes. Nevertheless, the method is fit-for-purpose, as the primary goal of our study was to profile and discover disease-specific trends rather than to achieve absolute quantification.

## Conclusions

This study is the first to monitor the abundance of α-synuclein peptides from the N-terminus and NAC-domain in plasma, revealing no detectable differential regulation of these domains among α-synucleinopathies via non-modified peptide measurements. Our findings provide a foundation for blood-based α-synuclein research, underscoring the need for assays that include post-translationally modified peptides to better understand the complexities of α-synucleinopathies.

## Data Availability

All data produced in the present study are available upon reasonable request to the authors

## List of abbreviations

AD: Alzheimer’s disease
AUC: Area under the curve
CMRR: Montpellier Memory Resources Center
CSF: Cerebrospinal fluid
DLB: Dementia with Lewy bodies
NAC: Non-amyloid-β component
NfL: Neurofilament light chain
MRM: Multiple Reaction Monitoring
MSA: Multiple system atrophy
MTBR: Microtubule-binding region
PD: Parkinson’s disease
PDD: Parkinson’s disease with dementia
pTau217: Tau protein with phosphorylation at the 217th amino acid residue
ROC: Receiver operating characteristic
SPE: Solid-phase extraction

## Declarations

### Ethical Approval and Consent to participate

Plasma was collected and stored at the Montpellier Neurobank (#DC-2008-417 of the certified NFS 96-900 CHU resource center BB-0033-00031). Authorization to handle personal data was granted by the French Data Protection Authority (CNIL). Participants gave their written consent to be enrolled in the study which was approved by the Montpellier University Hospital’s regional Ethics Committee.

### Consent for publication

Not applicable.

## Availability of data and materials

The dataset analyzed during the current study is available from the corresponding author on reasonable request.

## Competing interests Not applicable Funding

MLP received funding from Shimadzu corporation. PM has received funding from the European Union’s Horizon 2020 research and innovation programme under the Marie Skłodowska-Curie grant agreement No 860197 MIRIADE.

## Author contributions

MLP, JV, CH, SL conceptually designed the work. MLP developed the method. MLP and EM analyzed the patient samples. MLP performed the method validation. MLP processed the data. MLP and PM wrote the manuscript. PM performed data analysis. JV, SC, MV, SL, and CH reviewed the manuscript. All authors read and commented on the manuscript.

## Acknowledgements

We are grateful to all patients and their relatives for participating in this study.

Mass spectrometry experiments were carried out using the facilities of the Montpellier Proteomics Platform (PPM-PPC, BioCampus Montpellier). This article has been written in the context of the “innovation center” of Shimadzu Europa GmbH.

