## Supplementary material 1 for "Plasma α-synuclein domain profiles across α-synucleinopathies"

**Table 2.** Diagnostic accuracy of α-synuclein peptides and peptide combinations

| PD vs. Controls | n | AUC | 95% CI | Sensitivity | Specificity | Youden’s Index |
| --- | --- | --- | --- | --- | --- | --- |
| α-Syn 13-21 | 81/21 | 0.52 | [0.38 – 0.66] | 0.51 | 0.67 | 0.18 |
| α-Syn 24-32 | 81/21 | 0.56 | [0.42 – 0.70] | 0.62 | 0.62 | 0.24 |
| α-Syn 35-43 | 81/21 | 0.65 | [0.51 – 0.80] | 0.67 | 0.62 | 0.29 |
| α-Syn 61-80 | 81/21 | 0.5 | [0.36 – 0.64] | 0.59 | 0.57 | 0.16 |
| α-Syn 81-96 | 81/21 | 0.54 | [0.41 – 0.67] | 0.6 | 0.62 | 0.22 |
| α-Syn 13-21 + α-Syn 24-32 | 81/21 | 0.54 | [0.40 – 0.68] | 0.34 | 0.86 | 0.2 |
| α-Syn 13-21 + α-Syn 35-43 | 81/21 | 0.7 | [0.57 – 0.82] | 0.71 | 0.62 | 0.33 |
| α-Syn 13-21 + α-Syn 61-80 | 81/21 | 0.58 | [0.45 – 0.71] | 0.49 | 0.81 | 0.3 |
| α-Syn 13-21 + α-Syn 81-96 | 81/21 | 0.51 | [0.37 – 0.65] | 0.59 | 0.52 | 0.11 |
| α-Syn 24-32 + α-Syn 35-43 | 81/21 | 0.65 | [0.52 – 0.77] | 0.4 | 0.95 | 0.35 |
| α-Syn 24-32 + α-Syn 61-80 | 81/21 | 0.58 | [0.45 – 0.72] | 0.55 | 0.67 | 0.22 |
| α-Syn 24-32 + α-Syn 81-96 | 81/21 | 0.57 | [0.44 – 0.71] | 0.49 | 0.76 | 0.25 |
| α-Syn 35-43 + α-Syn 61-80 | 81/21 | 0.68 | [0.56 – 0.80] | 0.49 | 0.9 | 0.39 |
| α-Syn 35-43 + α-Syn 81-96 | 81/21 | 0.66 | [0.53 – 0.78] | 0.35 | 0.95 | 0.31 |
| α-Syn 61-80 + α-Syn 81-96 | 81/21 | 0.57 | [0.43 – 0.70] | 0.34 | 0.86 | 0.2 |
| PD vs. DLB | **n** | **AUC** | **95% CI** | **Sensitivity** | **Specificity** | **Youden’s Index** |
| α-Syn 13-21 | 81/32 | 0.55 | [0.44 – 0.67] | 0.28 | 0.91 | 0.19 |
| α-Syn 24-32 | 81/32 | 0.52 | [0.41 – 0.64] | 0.21 | 0.97 | 0.18 |
| α-Syn 35-43 | 81/32 | 0.50 | [0.39 – 0.62] | 0.37 | 0.75 | 0.12 |
| α-Syn 61-80 | 81/32 | 0.57 | [0.46 – 0.67] | 0.21 | 1 | 0.21 |
| α-Syn 81-96 | 81/32 | 0.54 | [0.43 – 0.65] | 0.22 | 0.94 | 0.16 |
| α-Syn 13-21 + α-Syn 24-32 | 81/32 | 0.53 | [0.41 – 0.66] | 0.78 | 0.41 | 0.19 |
| α-Syn 13-21 + α-Syn 35-43 | 81/32 | 0.57 | [0.45 – 0.69] | 0.62 | 0.53 | 0.15 |
| α-Syn 13-21 + α-Syn 61-80 | 81/32 | 0.5 | [0.37 – 0.62] | 0.57 | 0.5 | 0.07 |
| α-Syn 13-21 + α-Syn 81-96 | 81/32 | 0.52 | [0.40 – 0.64] | 0.46 | 0.66 | 0.12 |
| α-Syn 24-32 + α-Syn 35-43 | 81/32 | 0.56 | [0.44 – 0.68] | 0.7 | 0.53 | 0.23 |
| α-Syn 24-32 + α-Syn 61-80 | 81/32 | 0.57 | [0.45 – 0.69] | 0.74 | 0.47 | 0.21 |
| α-Syn 24-32 + α-Syn 81-96 | 81/32 | 0.54 | [0.41 – 0.66] | 0.44 | 0.69 | 0.13 |
| α-Syn 35-43 + α-Syn 61-80 | 81/32 | 0.58 | [0.46 – 0.71] | 0.67 | 0.56 | 0.23 |
| α-Syn 35-43 + α-Syn 81-96 | 81/32 | 0.56 | [0.44 – 0.68] | 0.72 | 0.47 | 0.19 |
| α-Syn 61-80 + α-Syn 81-96 | 81/32 | 0.53 | [0.40 – 0.66] | 0.68 | 0.5 | 0.18 |
| PD vs. MSA | **n** | **AUC** | **95% CI** | **Sensitivity** | **Specificity** | **Youden’s Index** |
| α-Syn 13-21 | 81/8 | 0.54 | [0.29 – 0.79] | 0.74 | 0.5 | 0.24 |
| α-Syn 24-32 | 81/8 | 0.54 | [0.28 – 0.79] | 0.73 | 0.5 | 0.23 |
| α-Syn 35-43 | 81/8 | 0.57 | [0.34 – 0.81] | 0.74 | 0.5 | 0.24 |
| α-Syn 61-80 | 81/8 | 0.52 | [0.27 – 0.78] | 0.72 | 0.5 | 0.22 |
| α-Syn 81-96 | 81/8 | 0.54 | [0.27 – 0.80] | 0.74 | 0.5 | 0.24 |
| α-Syn 13-21 + α-Syn 24-32 | 81/8 | 0.61 | [0.39 – 0.82] | 0.78 | 0.5 | 0.28 |
| α-Syn 13-21 + α-Syn 35-43 | 81/8 | 0.64 | [0.47 – 0.82] | 0.51 | 0.88 | 0.39 |
| α-Syn 13-21 + α-Syn 61-80 | 81/8 | 0.61 | [0.41 – 0.80] | 0.49 | 0.75 | 0.24 |
| α-Syn 13-21 + α-Syn 81-96 | 81/8 | 0.55 | [0.30 – 0.81] | 0.71 | 0.62 | 0.33 |
| α-Syn 24-32 + α-Syn 35-43 | 81/8 | 0.64 | [0.48 – 0.80] | 0.4 | 1 | 0.4 |
| α-Syn 24-32 + α-Syn 61-80 | 81/8 | 0.64 | [0.48 – 0.81] | 0.52 | 0.88 | 0.4 |
| α-Syn 24-32 + α-Syn 81-96 | 81/8 | 0.6 | [0.42 – 0.78] | 0.38 | 1 | 0.38 |
| α-Syn 35-43 + α-Syn 61-80 | 81/8 | 0.65 | [0.49 – 0.80] | 0.54 | 0.88 | 0.41 |
| α-Syn 35-43 + α-Syn 81-96 | 81/8 | 0.58 | [0.35 – 0.81] | 0.76 | 0.5 | 0.26 |
| α-Syn 61-80 + α-Syn 81-96 | 81/8 | 0.71 | [0.54 – 0.88] | 0.62 | 0.88 | 0.5 |
| DLB vs. Controls | **n** | **AUC** | **95% CI** | **Sensitivity** | **Specificity** | **Youden’s Index** |
| α-Syn 13-21 | 32/21 | 0.6 | [0.43 – 0.77] | 0.81 | 0.48 | 0.29 |
| α-Syn 24-32 | 32/21 | 0.63 | [0.46 – 0.79] | 0.78 | 0.57 | 0.35 |
| α-Syn 35-43 | 32/21 | 0.66 | [0.50 – 0.82] | 0.88 | 0.48 | 0.35 |
| α-Syn 61-80 | 32/21 | 0.58 | [0.41 – 0.75] | 0.59 | 0.67 | 0.26 |
| α-Syn 81-96 | 32/21 | 0.6 | [0.44 – 0.76] | 0.56 | 0.76 | 0.32 |
| α-Syn 13-21 + α-Syn 24-32 | 32/21 | 0.64 | [0.47 – 0.80] | 0.75 | 0.62 | 0.37 |
| α-Syn 13-21 + α-Syn 35-43 | 32/21 | 0.63 | [0.47 – 0.79] | 0.81 | 0.48 | 0.29 |
| α-Syn 13-21 + α-Syn 61-80 | 32/21 | 0.57 | [0.42 – 0.73] | 0.44 | 0.81 | 0.25 |
| α-Syn 13-21 + α-Syn 81-96 | 32/21 | 0.6 | [0.43 – 0.76] | 0.56 | 0.76 | 0.32 |
| α-Syn 24-32 + α-Syn 35-43 | 32/21 | 0.62 | [0.46 – 0.77] | 0.41 | 0.9 | 0.31 |
| α-Syn 24-32 + α-Syn 61-80 | 32/21 | 0.59 | [0.43 – 0.75] | 0.62 | 0.67 | 0.29 |
| α-Syn 24-32 + α-Syn 81-96 | 32/21 | 0.55 | [0.39 – 0.72] | 0.56 | 0.67 | 0.23 |
| α-Syn 35-43 + α-Syn 61-80 | 32/21 | 0.42 | [0.26 – 0.58] | 0.16 | 0.9 | 0.06 |
| α-Syn 35-43 + α-Syn 81-96 | 32/21 | 0.6 | [0.44 – 0.75] | 0.38 | 0.9 | 0.28 |
| α-Syn 61-80 + α-Syn 81-96 | 32/21 | 0.55 | [0.39 – 0.71] | 0.44 | 0.76 | 0.2 |
| DLB vs. MSA | **n** | **AUC** | **95% CI** | **Sensitivity** | **Specificity** | **Youden’s Index** |
| α-Syn 13-21 | 32/8 | 0.49 | [0.20 – 0.78] | 0.81 | 0.5 | 0.31 |
| α-Syn 24-32 | 32/8 | 0.51 | [0.21 – 0.81] | 0.84 | 0.5 | 0.34 |
| α-Syn 35-43 | 32/8 | 0.55 | [0.28 – 0.83] | 0.81 | 0.5 | 0.31 |
| α-Syn 61-80 | 32/8 | 0.49 | [0.19 – 0.79] | 0.81 | 0.5 | 0.31 |
| α-Syn 81-96 | 32/8 | 0.52 | [0.22 – 0.81] | 0.81 | 0.5 | 0.31 |
| α-Syn 13-21 + α-Syn 24-32 | 32/8 | 0.51 | [0.24 – 0.77] | 0.69 | 0.62 | 0.31 |
| α-Syn 13-21 + α-Syn 35-43 | 32/8 | 0.68 | [0.51 – 0.85] | 0.53 | 0.88 | 0.41 |
| α-Syn 13-21 + α-Syn 61-80 | 32/8 | 0.61 | [0.41 – 0.81] | 0.28 | 1 | 0.28 |
| α-Syn 13-21 + α-Syn 81-96 | 32/8 | 0.6 | [0.34 – 0.86] | 0.88 | 0.5 | 0.38 |
| α-Syn 24-32 + α-Syn 35-43 | 32/8 | 0.67 | [0.49 – 0.84] | 0.56 | 0.88 | 0.44 |
| α-Syn 24-32 + α-Syn 61-80 | 32/8 | 0.69 | [0.51 – 0.86] | 0.59 | 0.88 | 0.47 |
| α-Syn 24-32 + α-Syn 81-96 | 32/8 | 0.58 | [0.33 – 0.83] | 0.78 | 0.5 | 0.28 |
| α-Syn 35-43 + α-Syn 61-80 | 32/8 | 0.7 | [0.53 – 0.87] | 0.66 | 0.88 | 0.53 |
| α-Syn 35-43 + α-Syn 81-96 | 32/8 | 0.67 | [0.48 – 0.86] | 0.5 | 0.88 | 0.38 |
| α-Syn 61-80 + α-Syn 81-96 | 32/8 | 0.68 | [0.49 – 0.87] | 0.56 | 0.88 | 0.44 |
| MSA vs. Controls | **n** | **AUC** | **95% CI** | **Sensitivity** | **Specificity** | **Youden’s Index** |
| α-Syn 13-21 | 8/21 | 0.51 | [0.23 – 0.80] | 0.5 | 0.76 | 0.26 |
| α-Syn 24-32 | 8/21 | 0.56 | [0.27 – 0.85] | 0.62 | 0.71 | 0.34 |
| α-Syn 35-43 | 8/21 | 0.69 | [0.48 – 0.90] | 0.5 | 0.86 | 0.36 |
| α-Syn 61-80 | 8/21 | 0.51 | [0.23 – 0.79] | 0.62 | 0.67 | 0.29 |
| α-Syn 81-96 | 8/21 | 0.55 | [0.25 – 0.85] | 0.62 | 0.76 | 0.39 |
| α-Syn 13-21 + α-Syn 24-32 | 8/21 | 0.57 | [0.27 – 0.86] | 0.62 | 0.81 | 0.43 |
| α-Syn 13-21 + α-Syn 35-43 | 8/21 | 0.9 | [0.78 – 1.00] | 1 | 0.67 | 0.67 |
| α-Syn 13-21 + α-Syn 61-80 | 8/21 | 0.74 | [0.54 – 0.95] | 0.75 | 0.71 | 0.46 |
| α-Syn 13-21 + α-Syn 81-96 | 8/21 | 0.6 | [0.31 – 0.88] | 0.62 | 0.86 | 0.48 |
| α-Syn 24-32 + α-Syn 35-43 | 8/21 | 0.88 | [0.76 – 1.00] | 1 | 0.71 | 0.71 |
| α-Syn 24-32 + α-Syn 61-80 | 8/21 | 0.78 | [0.59 – 0.97] | 0.75 | 0.86 | 0.61 |
| α-Syn 24-32 + α-Syn 81-96 | 8/21 | 0.55 | [0.25 – 0.85] | 0.62 | 0.71 | 0.34 |
| α-Syn 35-43 + α-Syn 61-80 | 8/21 | 0.93 | [0.83 – 1.00] | 1 | 0.81 | 0.81 |
| α-Syn 35-43 + α-Syn 81-96 | 8/21 | 0.82 | [0.65 – 0.99] | 1 | 0.57 | 0.57 |
| α-Syn 61-80 + α-Syn 81-96 | 8/21 | 0.74 | [0.54 – 0.94] | 0.88 | 0.67 | 0.54 |
